# PTSD, Comorbidities, Gender, and Increased Risk of Cardiovascular Disease in a Large Military Cohort

**DOI:** 10.1101/2024.04.13.24305769

**Authors:** David S. Krantz, Frances H. Gabbay, Elizabeth A. Belleau, Pablo A. Aliaga, Gary H. Wynn, Murray B. Stein, Robert J. Ursano, James A. Naifeh

**Author notes:** **Corresponding Author:** David S. Krantz, Ph.D., Department of Medical and Clinical Psychology, Uniformed Services University of the Health Sciences, 4301 Jones Bridge Rd., Bethesda MD, 20814.

## Abstract

**Importance:** Posttraumatic stress disorder (PTSD) is a prevalent mental health problem that increases risk of cardiovascular disease (CVD). It is not known whether gender or comorbidities modify associations between PTSD and CVD.

**Objective:** To assess risk of hypertension and atherosclerotic CVD (ASCVD) associated with PTSD in a predominantly young military population, and determine if gender or PTSD comorbidities modify these associations.

**Design, setting, and participants:** Using administrative medical records, this longitudinal, retrospective cohort study assessed relationships of PTSD, gender, comorbidities (metabolic risk factors [MRF], behavioral risk factors [BRF], depression, and sleep disorders) to subsequent hypertension and ASCVD among 863,993 active-duty U.S. Army enlisted soldiers (86.2% male; 93.7% <age 40). Using discrete-time survival analysis, person-months with an initial hypertension diagnosis (*n*=49,656) were compared to an equal-probability control sample. Separate analyses compared person-months with ASCVD (n=2,427) to an equal-probability control sample.

**Main outcomes and measures:** ICD-9-CM diagnoses of hypertension, ASCVD (coronary artery disease, myocardial infarction, stroke, heart failure), PTSD, MRF (Type 2 diabetes, obesity), BRF (tobacco/alcohol use disorders), depression, and sleep disorders.

**Results:** PTSD was associated with subsequent hypertension (OR=3.0 [95% CI=2.9-3.1]), and ASCVD (OR=2.7 [95% CI=2.2-3.3]). These associations remained significant but were attenuated after adjusting for comorbidities and sociodemographic/service-related variables (Hypertension: OR=1.9 [95% CI=1.8-2.0]; ASCVD: OR=1.4 [95% CI=1.2-1.8]). For hypertension, gender and each comorbidity were significant explanatory variables in multivariable models, and there were significant PTSD interactions with gender, MRF, depression, and sleep disorders. Stratifying separately by gender and presence of each comorbidity, PTSD-hypertension associations were stronger among men, those without MRF, without depression, and without sleep disorders. Standardized risk estimates indicated that predicted hypertension rates for those with vs. without PTSD were higher for men, and for those with vs. without MRF, depression, and sleep disorders. For ASCVD, comorbidities, but not gender, were independent predictors, and associations between PTSD and ASCVD were not modified by gender or comorbidities.

**Conclusions and relevance:** PTSD and comorbidities are independent risk factors for hypertension and ASVD in younger individuals, and gender and comorbid conditions modify PTSD relationships with hypertension. These findings suggest that CVD preventive interventions address PTSD and medical and behavioral comorbidities.

## INTRODUCTION

Posttraumatic stress disorder (PTSD) is a stress-related disorder resulting from exposure to trauma and defined by the presence of intrusive thoughts, avoidance of trauma-related stimuli, negative alterations in cognitions and mood, and arousal and stress reactivity.^1^ Estimated PTSD lifetime prevalence is 6% in American adults, with a higher lifetime prevalence among military veterans.^2,3^

PTSD is also a risk factor for the development of cardiovascular disease (CVD), including hypertension and manifestations of atherosclerotic cardiovascular disease (ASCVD) such as coronary heart disease and stroke.^4–7^ The independent risk associated with PTSD is roughly comparable to that of other major CVD risk factors,^8–12^ and PTSD associations with CVD are present in both men^9,10,13^ and women.^14–16^ Since most studies have been conducted in older male veterans,^9,13^ it is unclear whether these associations are present in younger adults or whether there are gender differences in PTSD-CVD associations.^17,18^

PTSD is a systemic disorder that often co-occurs with medical and behavioral conditions that also are CVD risk factors.^19–24^ PTSD has a high comorbidity with depression,^25–27^ abdominal obesity,^28,29^ Type 2 diabetes mellitus (T2DM),^28^ tobacco^30^ and alcohol use disorders,^31^ and sleep disorders.^32–34^ Despite prior studies controlling for CVD risk factors,^9,11,35^ it is unclear whether comorbidities account for PTSD-related CVD risk,^19,36,37^ or whether comorbid conditions modify PTSD-related risk.^19^ Whether comorbidities contribute to, or modify, CVD risk associated with PTSD has implications for choosing clinical targets and treatments.

The present study examined relationships between PTSD and CVD incidence in a large cohort of male and female servicemembers predominantly under age 40,^38,39^ using electronic medical record diagnostic codes.^40^ Although incidence of hypertension and ASCVD each increase with age,^41^ hypertension is more likely to be clinically evident prior to middle age.^42,43^ Therefore, we separately examined hypertension and ASCVD. We hypothesized that PTSD would be independently associated with subsequent hypertension and ASCVD in this younger population, and that controlling for comorbidities would attenuate but not eliminate this relationship. We also determined whether PTSD-associated risk is modified by gender or by presence of comorbidities.

## METHODS

### Sample

This longitudinal, retrospective cohort study used data from the Army Study to Assess Risk and Resilience in Servicemembers (Army STARRS) Historical Administrative Data Study (HADS),^39^ which integrates all Army and Department of Defense data systems documenting medical outcomes. The HADS includes individual-level person-month records for all soldiers (n≈1.66 million) on active duty at any point between January 1, 2004, and December 31, 2009, with health data available since 2000. We excluded officers and National Guard and Reserve members and used administrative records from all 863,993 Regular Army enlisted soldiers on active duty between 2004-2009 to select two independent samples: one for hypertension (with no prior or concurrent ASCVD diagnosis) and one for ASCVD (with or without prior/concurrent hypertension). This study was approved by the Uniformed Services University Institutional Review Board and follows Strengthening the Reporting of Observational Studies in Epidemiology (STROBE) cohort study guidelines.

Data were analyzed using a discrete-time survival framework with person-month the unit of analysis^44,45^ and each month of a soldier’s career represented as a separate record. To form a sample with hypertension as the sole CVD diagnosis (Hypertension Only), we identified all soldiers receiving a hypertension diagnosis without prior/concurrent ASCVD, selecting for analysis the person-month with first hypertension diagnosis (n=49,656 cases). To reduce computation demands, an equal-probability control sample was selected to be representative of all remaining person-months in the Regular Army enlisted population (n=1,052,151 unweighted person-months). Person-month data were first stratified by gender and service-related variables (rank, time in service, deployment status), then control person-months were selected on a 20:1 ratio to cases and given a weight of 29.7 (inverse probability of selection) to adjust for under-sampling. A separate sample of ASCVD cases (with or without prior/concurrent hypertension diagnosis) was selected using an identical process, resulting in 2,427 ASCVD cases (person-months receiving initial ASCVD diagnosis), and 87,722 control person-months, the latter assigned a weight of 362.1 to adjust for under-sampling (Figure 1).

**Figure 1.**
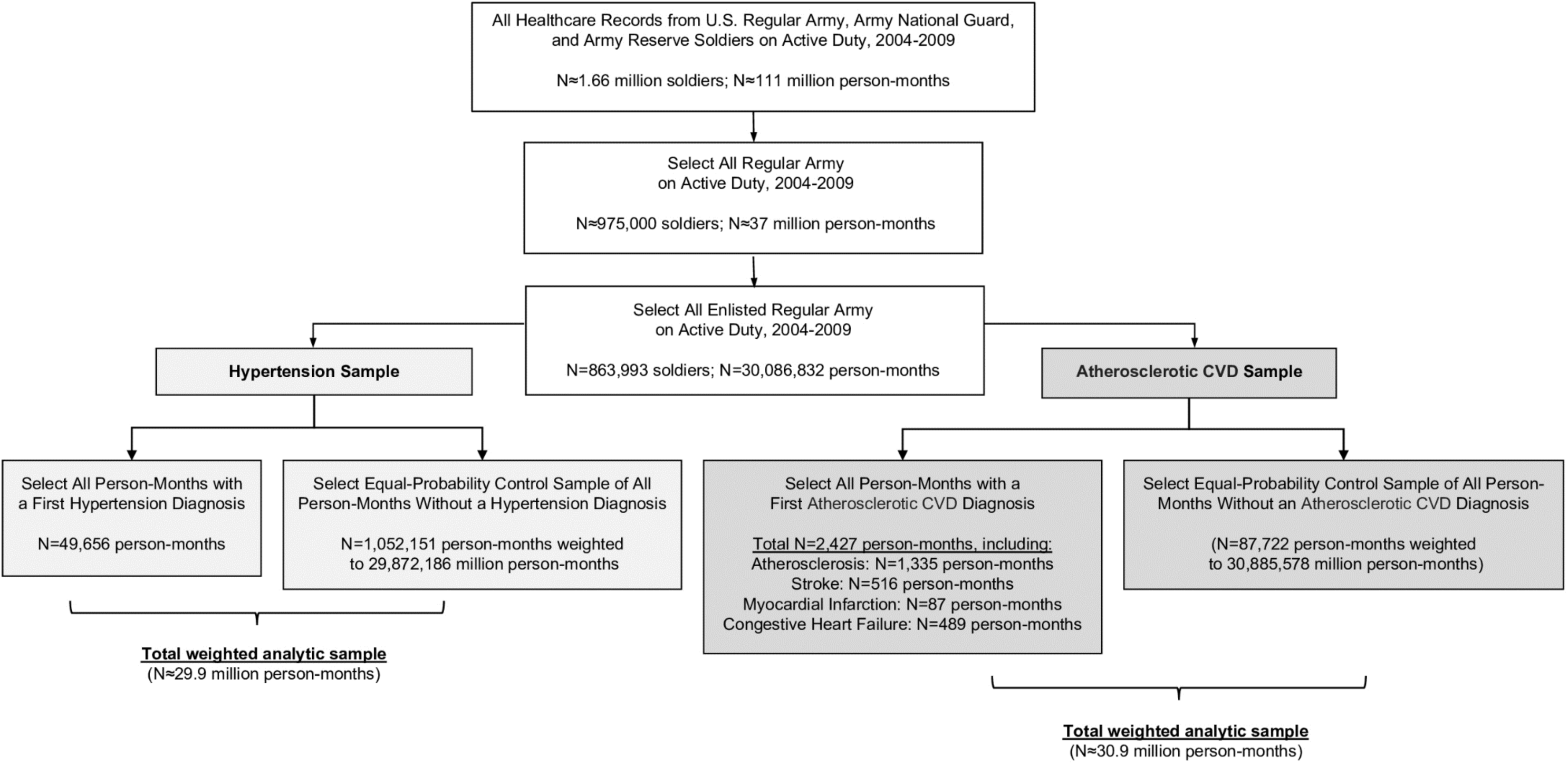
Flowchart representing the selection of two samples of Regular Army enlisted soldiers based on ICD-9-CM CVD diagnoses: (1) all person-months with a first hypertension diagnosis (without prior/concurrent ASCVD), and an equal-probability sample of control person-months (assigned a weight of 29.7 to adjust for under-sampling); and (2) all person-months with a first ASCVD diagnosis (with or without prior/concurrent hypertension diagnosis) and an equal-probability sample of control person-months (assigned a weight of 362.1). ASCVD=atherosclerotic cardiovascular disease; CVD=cardiovascular disease; ICD-9-CM=International Classification of Diseases, Ninth Revision, Clinical Modification.

### Measures

Cardiovascular disease (CVD) was identified using *International Classification of Diseases, Ninth Revision, Clinical Modification* (*ICD-9-CM*) diagnostic codes^46^ from the Military Health System (MHS) Data Repository and two other MHS databases, providing healthcare encounter information from military and civilian treatment facilities, deployment, and evacuation activities (Supplement, eTable 1). ICD-9-CM codes for CVD (Supplement, eTable 2) were classified as Hypertension Only or ASCVD (atherosclerosis, myocardial infarction, stroke, and/or congestive heart failure). In each sample, cases were identified based on the first person-month with the CVD diagnosis of interest (Figure 1). Person-month records were right censored after diagnosis.

ICD-9-CM codes were utilized to identify PTSD, depression, and sleep disorders, and create variables for metabolic risk factors (MRF; T2DM and/or obesity), and behavioral risk factors (BRF; tobacco and/or alcohol use disorder) (Supplement, eTable 3). PTSD diagnosis was examined only if documented *prior to* the first person-month with CVD diagnosis (for cases), or in the sampled person-month (for controls). Gender, race/ethnicity, age, and service-related variables were derived from administrative records (Supplement, eTable1).

### Statistical Analyses

Using SAS version 9.4, person-month data were analyzed using discrete-time survival analysis with a logit link function.^44,45^ Associations of PTSD with subsequent hypertension were examined in a univariable logistic regression model, followed by a multivariable model adjusting for sociodemographic and service-related variables, MRF, BRF, depression, and sleep disorders. To determine whether PTSD associations with hypertension differed by gender or presence/absence of comorbidities, two-way interactions of PTSD with gender, MRF, BRF, depression, and sleep disorders were examined in separate covariate-adjusted models. For significant interactions, the sample was stratified by presence/absence of each comorbidity, and the association between PTSD and CVD outcome was examined in a multivariable model within each stratum. The same analytic approach was used to examine associations of PTSD with ASCVD diagnosis.

Logistic regression coefficients were exponentiated to obtain odds ratios (ORs) and 95% CIs. Models included a month/year dummy predictor to control for secular trends; coefficients therefore represent averaged within-month associations assuming no time-varying effects.

Expected hypertension and ASCVD rates for those with and without PTSD were derived using full multivariable model coefficients to calculate standardized risk estimates (SREs) for PTSD (yes/no). All SREs are expressed as estimated number of individuals with each CVD endpoint per 1,000 person-years. To reduce Type I error, statistical significance of each Wald χ^2^ in the logistic regression models was based on a sample-size-adjusted alpha level^47^ requiring p<0.00048 for hypertension analyses and p<0.00167 for ASCVD analyses.

## RESULTS

### Sample Characteristics

In the total population, there were 49,656 cases of hypertension only (5.7%) and 2,427 ASCVD cases (0.3%) (*n*=853 ASCVD with prior/concurrent hypertension; *n*=1,574 without hypertension). The weighted hypertension case-control sample (Table 1) was predominantly male; non-Hispanic White; and young (median age=25 years [IQR 22-31]; 93.7% <age 40). A minority had diagnoses of PTSD (1.5%), MRF (4.1%), BRF (11.8%), depression (6.2%), and/or sleep disorders (3.3%). The weighted ASCVD case-control sample (Table 1) was predominantly male; non-Hispanic White; and young (median age=25 years [IQR 22-32], 93.2% <age 40). A minority had diagnoses of PTSD (1.6%), MRF (4.5%), BRF (12.0%), depression (6.4%), and/or sleep disorders (3.6%). Among hypertension cases, 2,201 (4.4%) had prior PTSD (median 10 [IQR 5-20] months between PTSD and hypertension diagnoses). Among ASCVD cases, 107 (4.4%) had prior PTSD (median 10 [IQR 5-29] months between PTSD and ASCVD).

**Table 1.**
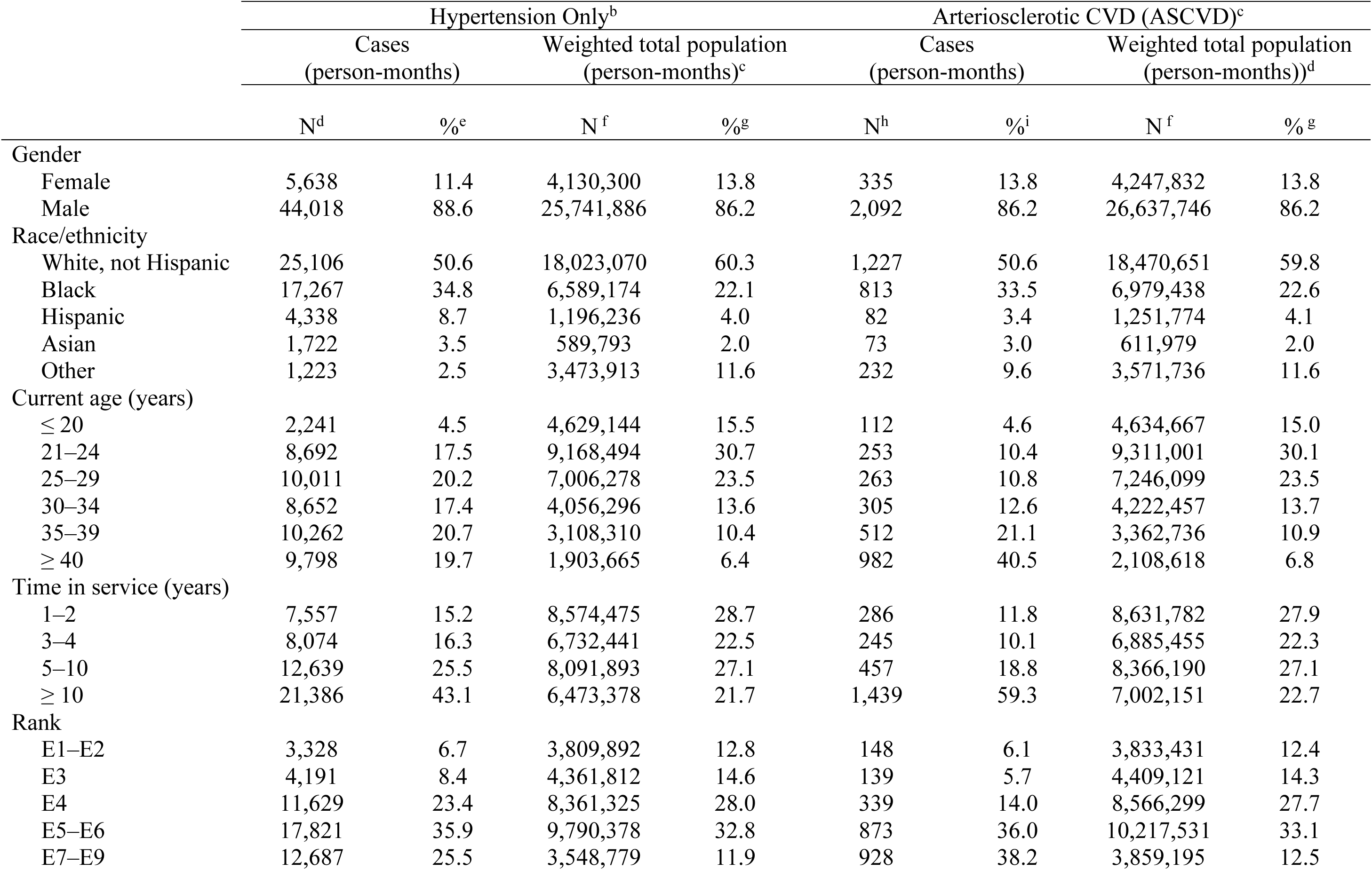
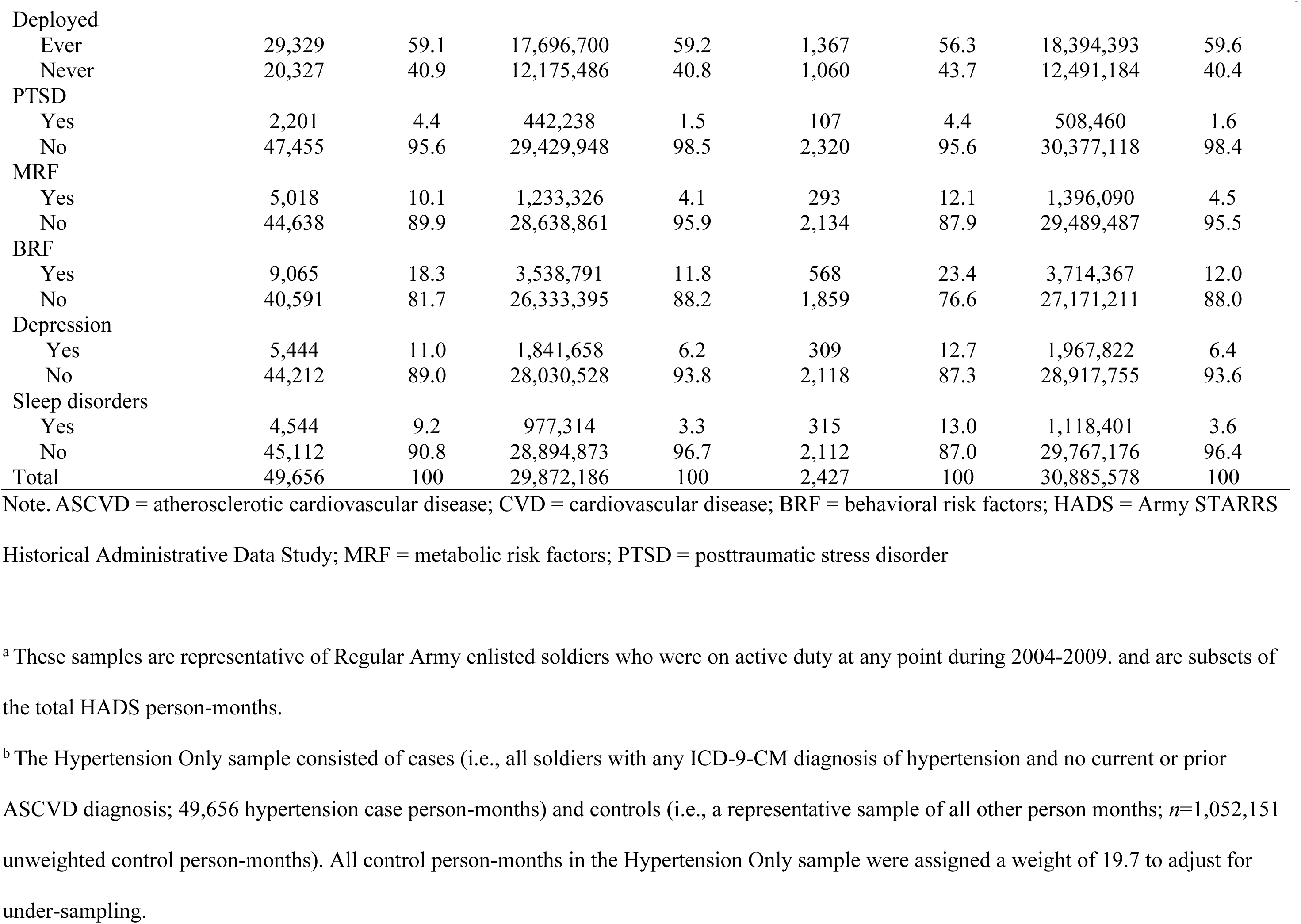
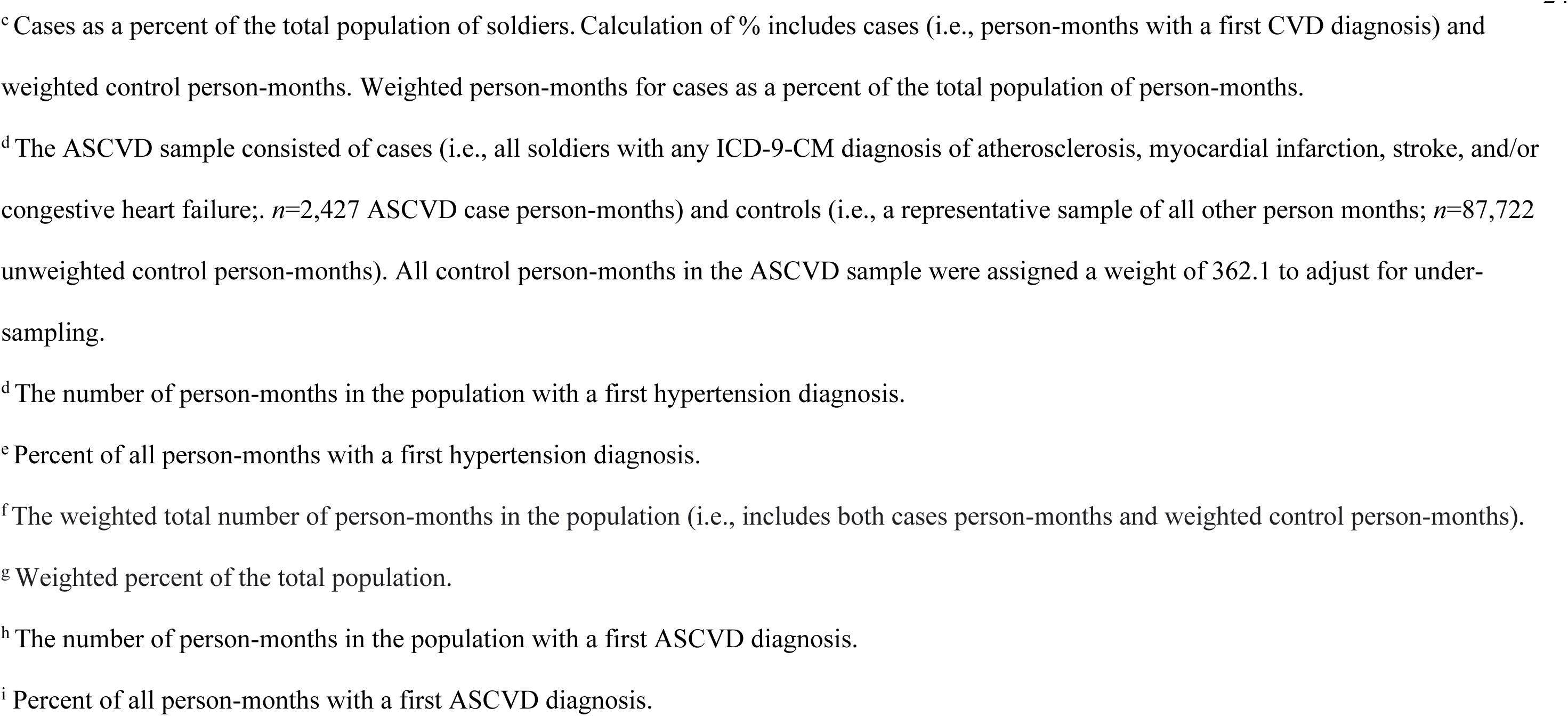
Distribution of Sociodemographic/Service-Related Variables and PTSD and Comorbidity Diagnoses Among Regular Army Enlisted Soldiers During 2004–2009^a^

### Association of PTSD with Hypertension

Controlling only for historical time, PTSD was associated with increased odds of developing hypertension (OR= 3.0 [95% CI=2.86–3.12]). Hypertension crude rates among those with vs. without PTSD were 59.7 vs. 19.4 per 1,000 person-years. Though attenuated, this association persisted after adjusting for MRF, BRF, depression, sleep disorders, and sociodemographic/service-related variables (OR=1.9 [95% CI=1.81–1.99]). Based on this full model, SREs for hypertension were 31.2 vs. 16.3 per 1,000 person-years among those with vs. without PTSD.

In the full model, there were also significant independent relationships between each risk variable and hypertension (Table 2). Odds of developing hypertension were higher among those with MRF (OR=1.8 [95% CI=1.75–1.86]), BRF (OR=1.3 [95% CI=1.30–1.36]), depression (OR=1.3 [95% CI=1.23–1.31]), and sleep disorders (OR=1.5 [95% CI=1.48–1.58]), and lower among women than men (OR=0.7 [95% CI=0.64–0.68]).

**Table 2.**
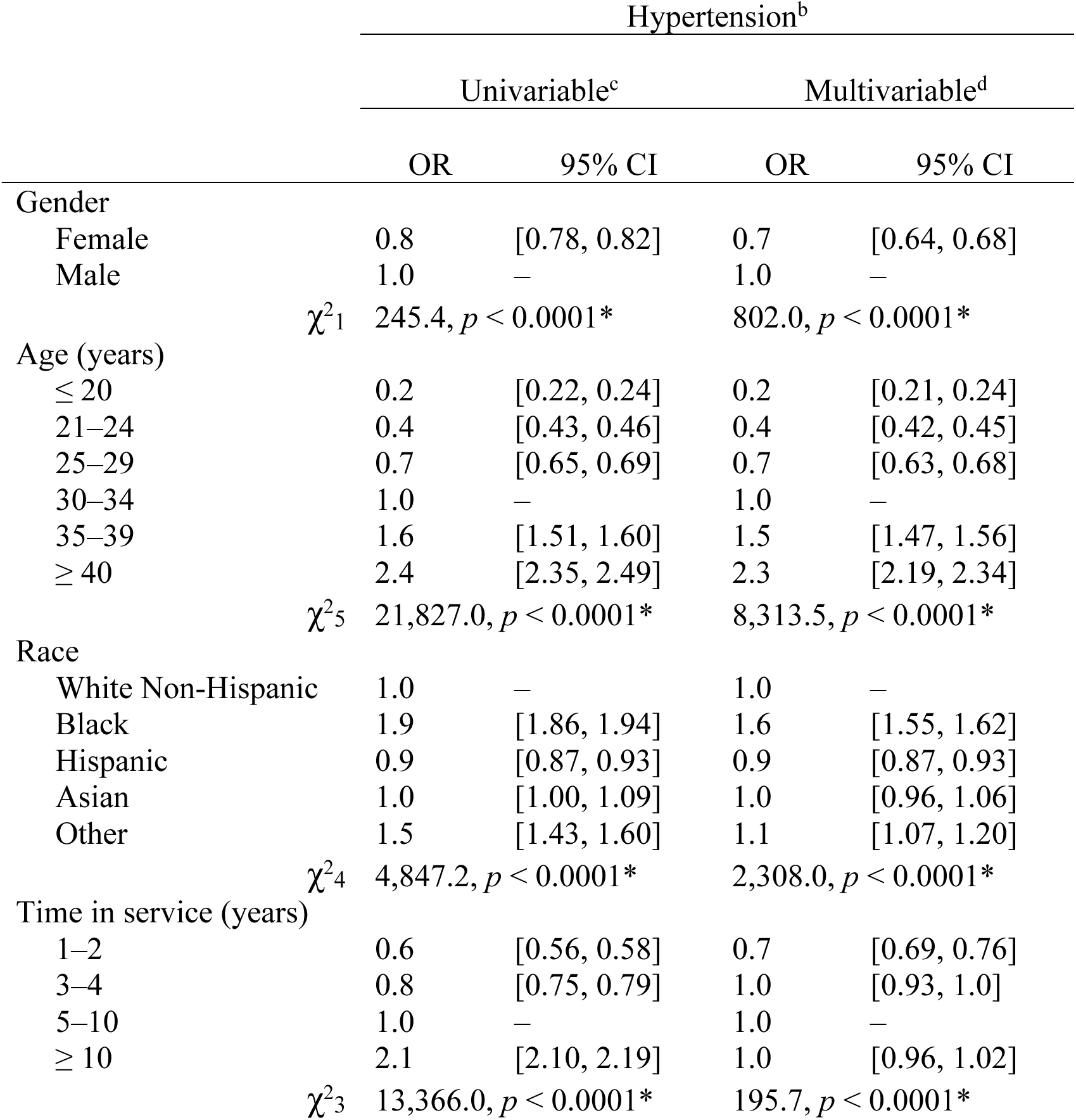
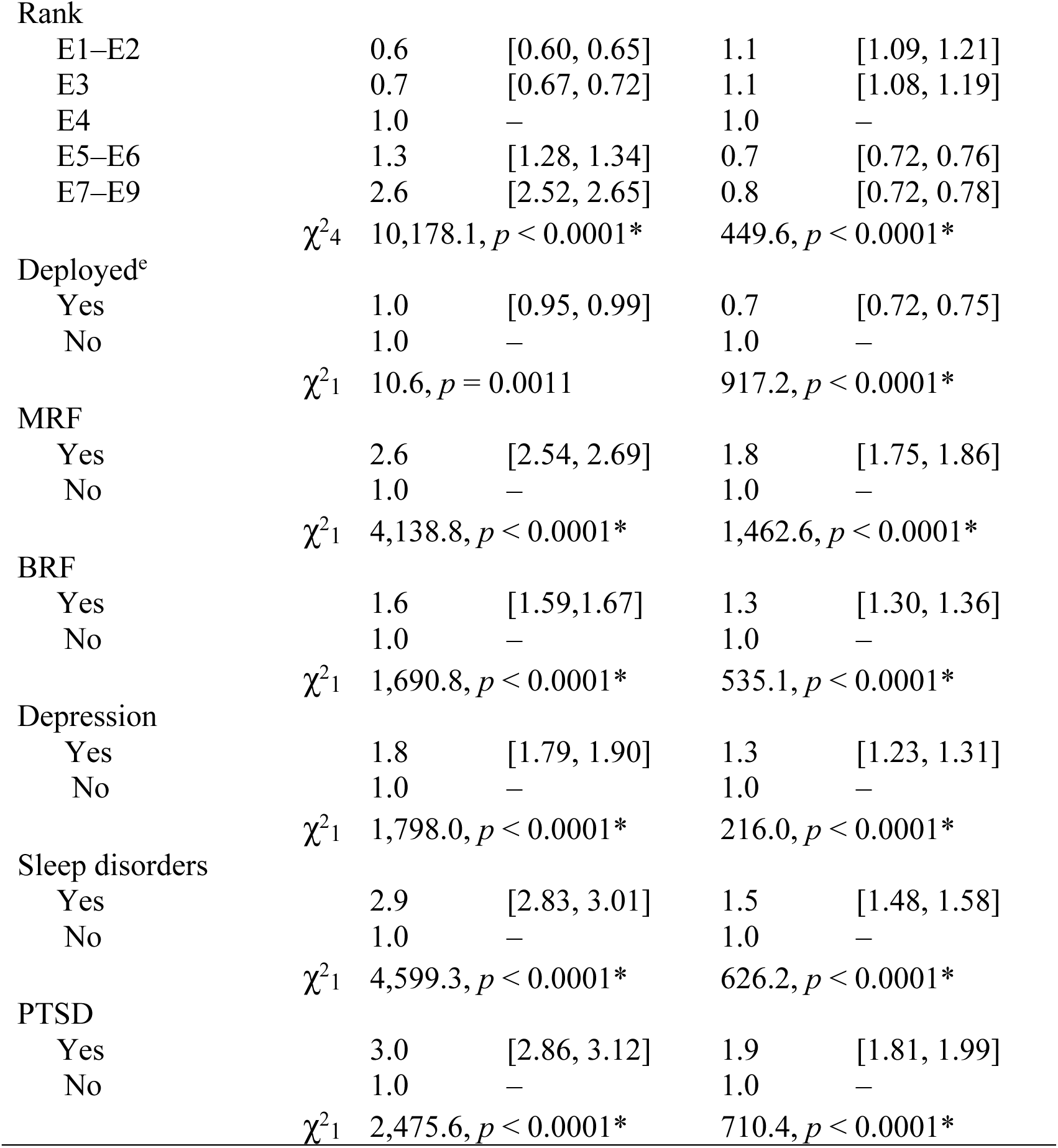
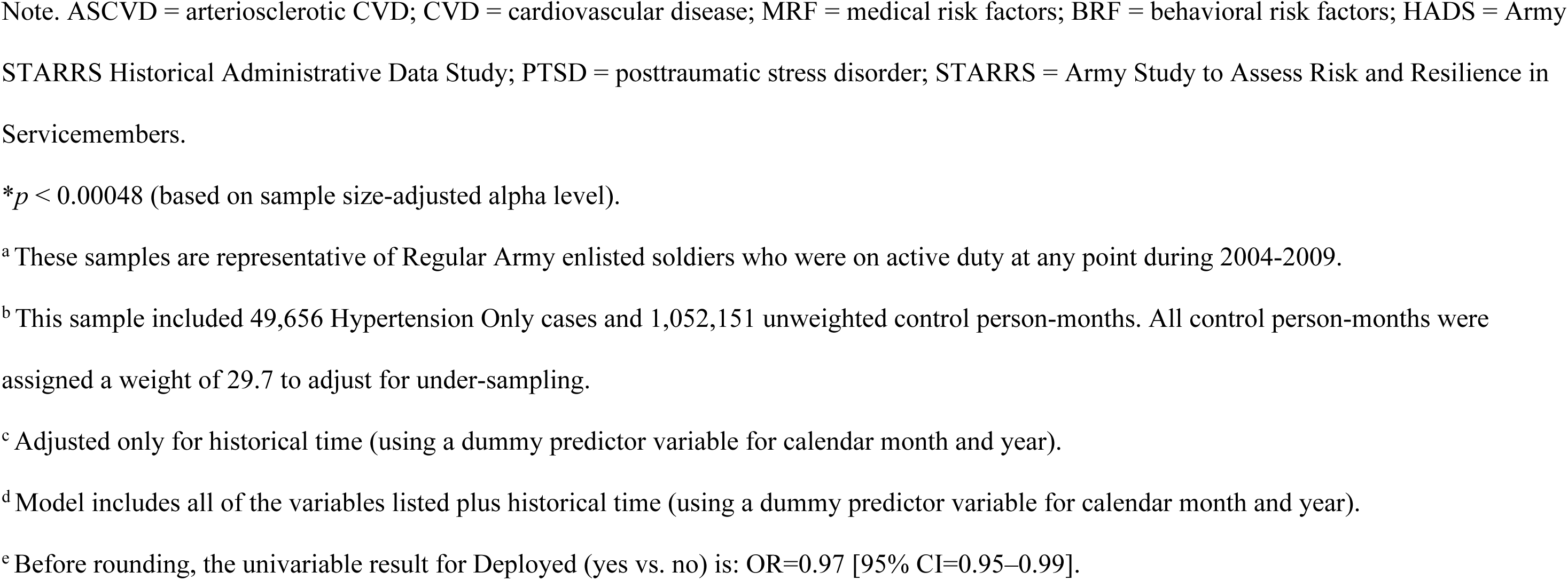
Univariable and Multivariable Associations of First PTSD Diagnosis with Subsequent Hypertension Diagnosis among Regular Army Enlisted

Modifiers of the PTSD-hypertension relationship were examined using separate multivariable models. There were significant two-way interactions between PTSD and gender (χ^2^1=43.9, *p*<0.0001), MRF (χ^2^1=21.3, *p*<0.0001), depression (χ^2^1=18.9, *p*<0.0001), and sleep disorders (χ^2^1=17.7, *p*<0.0001). The interaction with BRF was nonsignificant (χ^2^1=2.7, *p*=0.098).

To understand significant interactions, the sample was stratified separately by gender and each comorbidity, and SREs were calculated within each stratum for individuals with vs. without PTSD. The multivariable association of PTSD with hypertension was significant among men (OR=1.9 [95% CI=1.84–2.03]), and significant but smaller among women (OR=1.4 [95% CI=1.18–1.64]). Among men, standardized hypertension rates were higher for soldiers with vs. without PTSD (SREs=38.7 vs. 20.1 per 1,000 person-years, respectively), as they were among women (SREs=22.6 vs. 16.2) (Figure 2a). Overall, SREs were lower among women relative to men (Supplement, eTables 4-6).

**Figure 2.**
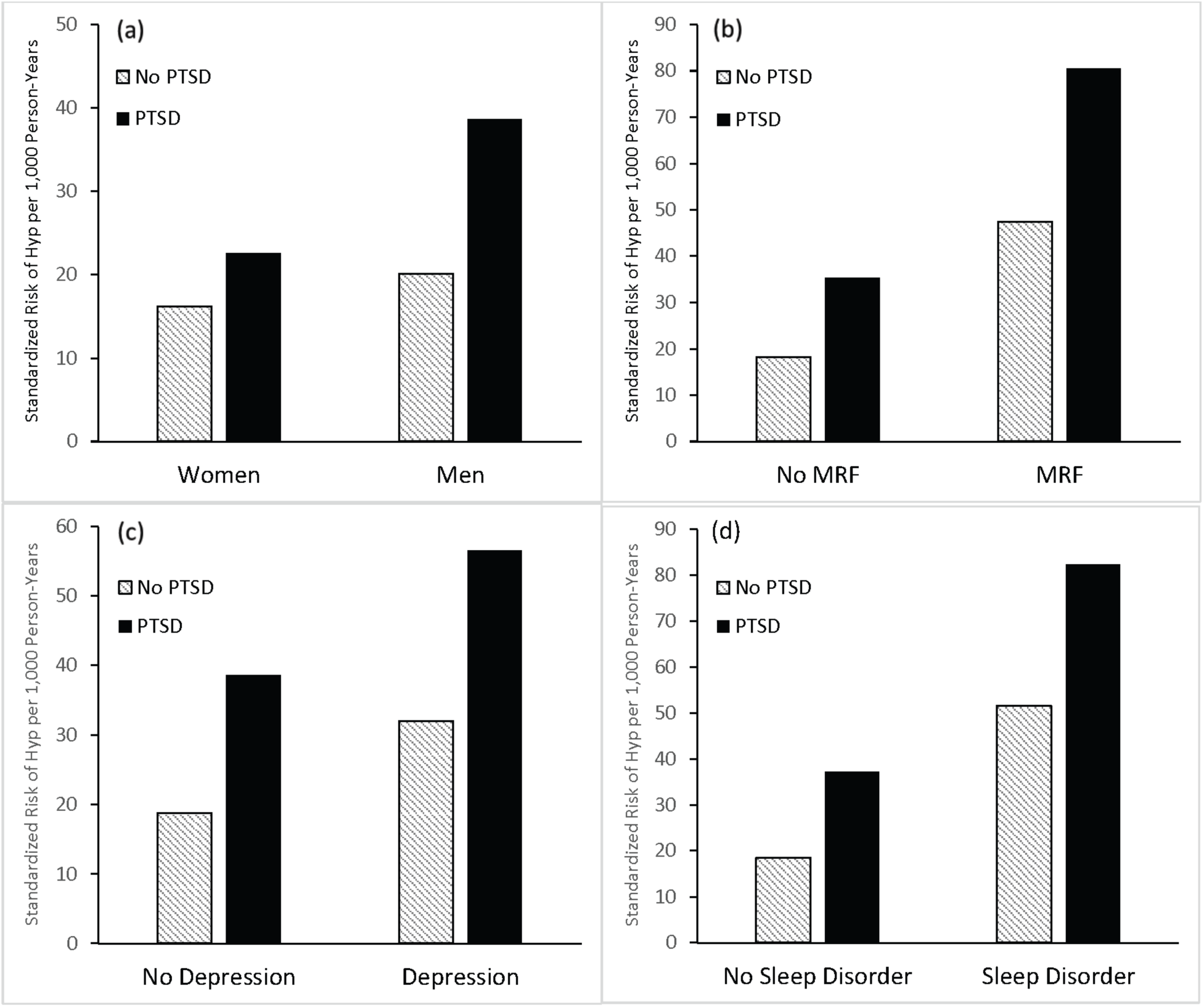
Standardized risk estimates of hypertension among individuals with and without posttraumatic stress disorder (PTSD) based on multivariable logistic regression analyses conducted within strata defined by gender and presence of other conditions and disorders. Panel (a): Gender (men vs. women); Panel (b): individuals with vs. without a metabolic risk factor (MRF; T2DM and/or obesity); Panel (c): individuals vs. without depression; and Panel (d): individuals with vs. without a sleep disorder.

Stratifying by MRF revealed a slightly stronger association between PTSD and hypertension incidence among individuals without (OR=1.9 [95% CI=1.84–2.04]) vs. with MRF (OR=1.7 [95% CI=1.51–1.92]). Among soldiers with MRF, standardized hypertension rates were higher among those with vs. without PTSD (SREs=80.5 vs. 47.4), as they were among soldiers without MRF (SREs=35. 3 vs 18.3). Overall, standardized rates of hypertension were lower among individuals without MRF (Figure 2b).

For depression, the association between PTSD and hypertension was also slightly stronger among individuals without (OR=2.1 [95% CI=1.95-2.22]) vs. with depression (OR=1.8 [95% CI=1.65-1.90]). Among soldiers with depression, standardized hypertension rates were higher among those with vs. without PTSD (SREs=56.6 vs. 32.0), as they were among soldiers without depression (SREs=38.7 vs. 18.7). Overall, standardized rates of hypertension were lower among individuals without depression (Figure 2c).

PTSD was more strongly associated with hypertension among individuals without (OR=2.0 [95% CI 1.91-2.15]) vs. with sleep disorders (OR=1.6 [95% CI 1.48-1.73]). Among soldiers with sleep disorders, standardized hypertension rates were higher among those with PTSD (SREs=82.3 vs. 51.6), as they were among soldiers without a sleep disorder (SRE=37.3 vs. 18.5). Overall, standardized rates of hypertension were lower among individuals without sleep disorders (Figure 2d).

The pattern of results indicates that standardized risk of hypertension is highest when both PTSD and a comorbidity are present and lowest when both are absent (Figure 2).

### Association of PTSD with ASCVD

PTSD was associated with increased odds of ASCVD, controlling only for historical time (OR=2.7 [95% CI=2.25–3.32]). ASCVD crude rates were 2.5 vs. 0.9 per 1,000 person-years for those with vs. without PTSD. Though attenuated, the association persisted after adjusting for MRF, BRF, depression, sleep disorders, and sociodemographic/service-related variables (OR=1.4 [95% CI=1.17–1.79]). Based on this multivariable model, SREs for ASCVD were 1.1 vs. 0.8 for those with vs. without PTSD.

In full multivariable models (Table 3), odds of developing ASCVD were also higher among those with MRF (OR=1.6 [95% CI=1.45–1.87]), BRF (OR=1.9 [95% CI=1.69–2.06]), depression (OR=1.3 [95% CI=1.14–1.49]), and sleep disorders (OR=1.6 [95% CI=1.44–1.86]). The association between gender and ASCVD was nonsignificant using the adjusted alpha level (*χ^2^1*=4.2, *p*=0.0393). Similarly, none of the two-way interactions between PTSD and gender or comorbidity was significant.

**Table 3.**
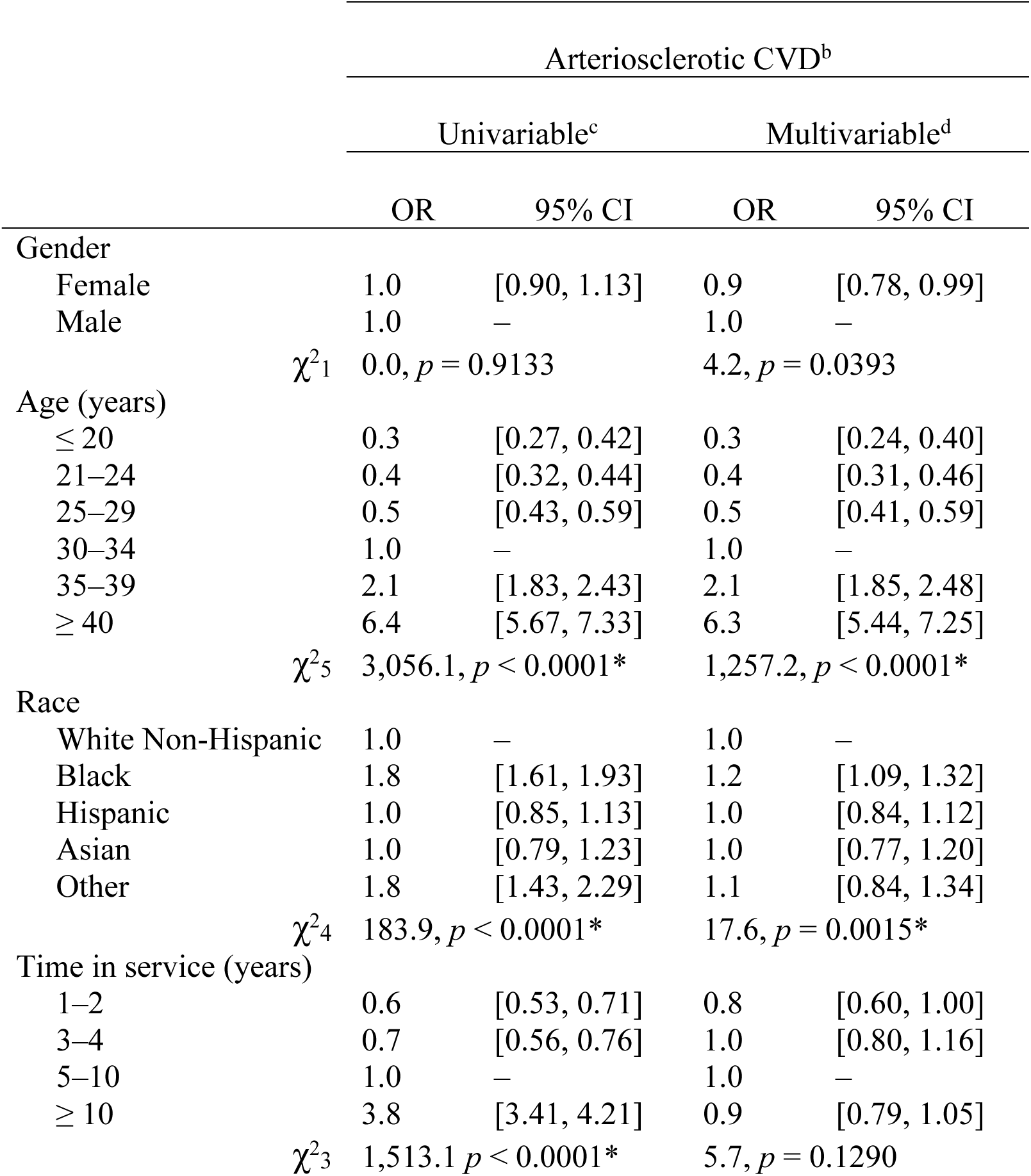
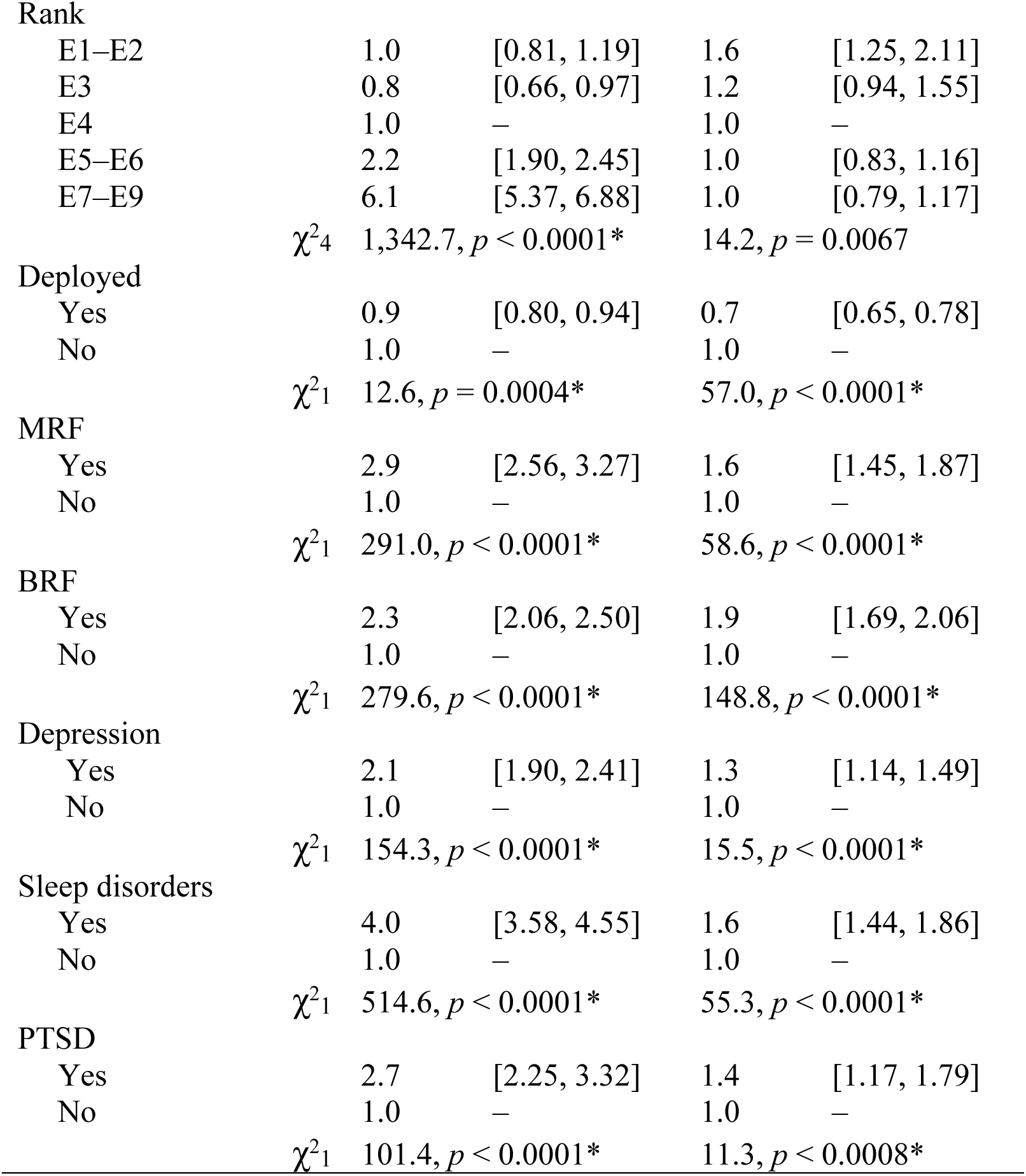
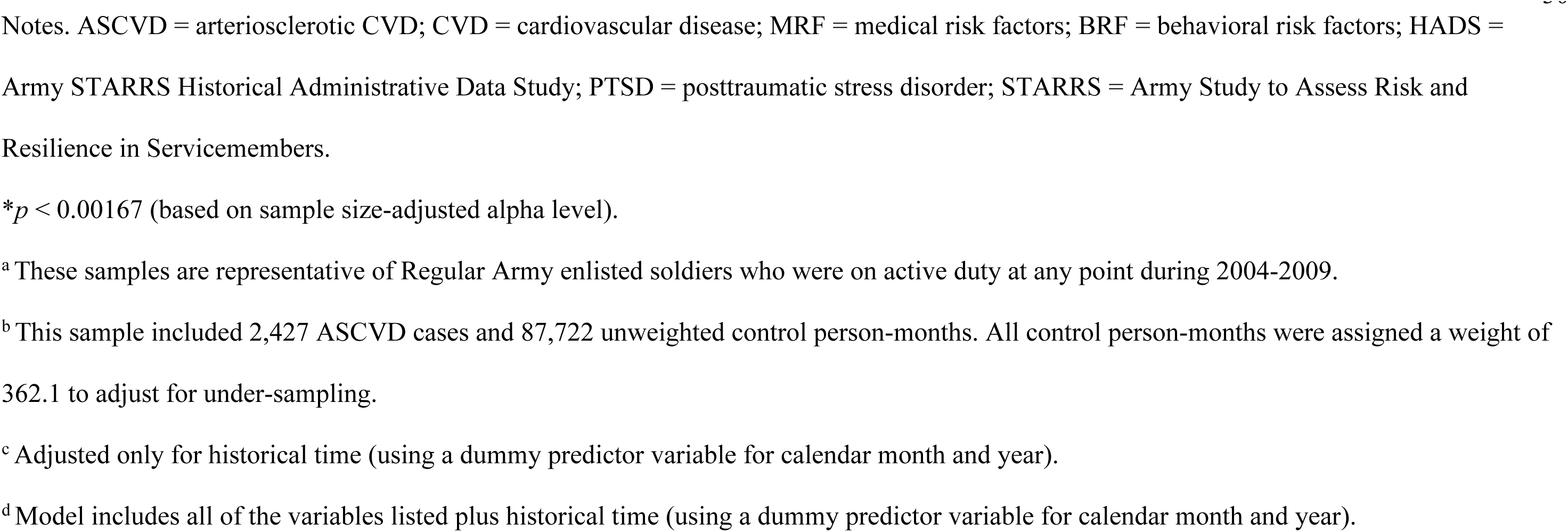
Univariable and Multivariable Associations of First PTSD Diagnosis with Subsequent ASCVD Diagnosis among Regular Army Enlisted Soldiers^a^

|  | Arteriosclerotic CVD<sup>b</sup> |  |  |  |
| --- | --- | --- | --- | --- |
|  | Univariable<sup>c</sup> |  | Multivariable<sup>d</sup> |  |
|  | OR | 95% CI | OR | 95% CI |
| Gender |  |  |  |  |
| Female | 1.0 | [0.90, 1.13] | 0.9 | [0.78, 0.99] |
| Male | 1.0 | — | 1.0 | — |
| | $\chi^2_1$ | 0.0, $p = 0.9133$ | | 4.2, $p = 0.0393$ |
| Age (years) |  |  |  |  |
| $\leq 20$ | 0.3 | [0.27, 0.42] | 0.3 | [0.24, 0.40] |
| 21–24 | 0.4 | [0.32, 0.44] | 0.4 | [0.31, 0.46] |
| 25–29 | 0.5 | [0.43, 0.59] | 0.5 | [0.41, 0.59] |
| 30–34 | 1.0 | — | 1.0 | — |
| 35–39 | 2.1 | [1.83, 2.43] | 2.1 | [1.85, 2.48] |
| $\geq 40$ | 6.4 | [5.67, 7.33] | 6.3 | [5.44, 7.25] |
| | $\chi^2_5$ | 3,056.1, $p < 0.0001^*$ | | 1,257.2, $p < 0.0001^*$ |
| Race |  |  |  |  |
| White Non-Hispanic | 1.0 | — | 1.0 | — |
| Black | 1.8 | [1.61, 1.93] | 1.2 | [1.09, 1.32] |
| Hispanic | 1.0 | [0.85, 1.13] | 1.0 | [0.84, 1.12] |
| Asian | 1.0 | [0.79, 1.23] | 1.0 | [0.77, 1.20] |
| Other | 1.8 | [1.43, 2.29] | 1.1 | [0.84, 1.34] |
| | $\chi^2_4$ | 183.9, $p < 0.0001^*$ | | 17.6, $p = 0.0015^*$ |
| Time in service (years) |  |  |  |  |
| 1–2 | 0.6 | [0.53, 0.71] | 0.8 | [0.60, 1.00] |
| 3–4 | 0.7 | [0.56, 0.76] | 1.0 | [0.80, 1.16] |
| 5–10 | 1.0 | — | 1.0 | — |
| $\geq 10$ | 3.8 | [3.41, 4.21] | 0.9 | [0.79, 1.05] |
| | $\chi^2_3$ | 1,513.1 $p < 0.0001^*$ | | 5.7, $p = 0.1290$ |
| Rank |  |  |  |  |
| E1–E2 | 1.0 | [0.81, 1.19] | 1.6 | [1.25, 2.11] |
| E3 | 0.8 | [0.66, 0.97] | 1.2 | [0.94, 1.55] |
| E4 | 1.0 | – | 1.0 | – |
| E5–E6 | 2.2 | [1.90, 2.45] | 1.0 | [0.83, 1.16] |
| E7–E9 | 6.1 | [5.37, 6.88] | 1.0 | [0.79, 1.17] |
| | $\chi^2_4$ | 1,342.7, $p < 0.0001^*$ | 14.2, $p = 0.0067$ | |
| Deployed |  |  |  |  |
| Yes | 0.9 | [0.80, 0.94] | 0.7 | [0.65, 0.78] |
| No | 1.0 | – | 1.0 | – |
| | $\chi^2_1$ | 12.6, $p = 0.0004^*$ | 57.0, $p < 0.0001^*$ | |
| MRF |  |  |  |  |
| Yes | 2.9 | [2.56, 3.27] | 1.6 | [1.45, 1.87] |
| No | 1.0 | – | 1.0 | – |
| | $\chi^2_1$ | 291.0, $p < 0.0001^*$ | 58.6, $p < 0.0001^*$ | |
| BRF |  |  |  |  |
| Yes | 2.3 | [2.06, 2.50] | 1.9 | [1.69, 2.06] |
| No | 1.0 | – | 1.0 | – |
| | $\chi^2_1$ | 279.6, $p < 0.0001^*$ | 148.8, $p < 0.0001^*$ | |
| Depression |  |  |  |  |
| Yes | 2.1 | [1.90, 2.41] | 1.3 | [1.14, 1.49] |
| No | 1.0 | – | 1.0 | – |
| | $\chi^2_1$ | 154.3, $p < 0.0001^*$ | 15.5, $p < 0.0001^*$ | |
| Sleep disorders |  |  |  |  |
| Yes | 4.0 | [3.58, 4.55] | 1.6 | [1.44, 1.86] |
| No | 1.0 | – | 1.0 | – |
| | $\chi^2_1$ | 514.6, $p < 0.0001^*$ | 55.3, $p < 0.0001^*$ | |
| PTSD |  |  |  |  |
| Yes | 2.7 | [2.25, 3.32] | 1.4 | [1.17, 1.79] |
| No | 1.0 | – | 1.0 | – |
| | $\chi^2_1$ | 101.4, $p < 0.0001^*$ | 11.3, $p < 0.0008^*$ | |

Notes. ASCVD = arteriosclerotic CVD; CVD = cardiovascular disease; MRF = medical risk factors; BRF = behavioral risk factors; HADS = Army STARRS Historical Administrative Data Study; PTSD = posttraumatic stress disorder; STARRS = Army Study to Assess Risk and Resilience in Servicemembers.
\* $p < 0.00167$ (based on sample size-adjusted alpha level).
<sup>a</sup> These samples are representative of Regular Army enlisted soldiers who were on active duty at any point during 2004-2009.
<sup>b</sup> This sample included 2,427 ASCVD cases and 87,722 unweighted control person-months. All control person-months were assigned a weight of 362.1 to adjust for under-sampling.
<sup>c</sup> Adjusted only for historical time (using a dummy predictor variable for calendar month and year).
<sup>d</sup> Model includes all of the variables listed plus historical time (using a dummy predictor variable for calendar month and year).

In a sensitivity analysis adding hypertension to the multivariable ASCVD model, hypertension was significant (OR=3.3 [95% CI=3.00–3.63]), but the PTSD-ASCVD association was no longer significant (OR=1.4 [95% CI=1.10–1.68]; *χ^2^1*=7.8, *p*=.0052) (Supplement, eTable 7).

## DISCUSSION

This study demonstrates that PTSD is associated with increased risk of both incident hypertension and ASCVD in servicemembers predominantly <40 years old. Comparing those with vs. without PTSD, crude rates (per 1,000 person-years) were 59.7 vs. 19.4 for hypertension and 2.5 vs. 0.9 for ASCVD. PTSD, MRF (obesity and/or T2DM), BRF (tobacco and/or alcohol use disorders), depression, and sleep disorders each independently increased risk of hypertension and ASCVD. Interactions with PTSD indicate further that gender, MRF, depression, and sleep disorders each moderate the relationship between PTSD and hypertension, but not ASCVD.

### PTSD and Gender

Consistent with prior research^4–12^ PTSD was associated with risk of subsequent CVD among both men and women.^9,10,14–16,48^ In this relatively young cohort, this risk was higher among men than women, and the association of PTSD with hypertension (but not ASCVD) was stronger among men. One study demonstrated stronger relationships between PTSD and ischemic stroke in men than women;^5^ however, prior studies have not specifically examined interactions between PTSD and gender for hypertension or ASCVD.

PTSD prevalence is higher among women than men.^25^ CVD risk increases across the lifespan in both genders,^43^ but is relatively low in premenopausal women.^49,50^ In our predominantly premenopausal female sample, gender differences in PTSD associations with hypertension may be partially explained by protective effects of estrogen^51^ and/or lower CVD incidence among younger women.^50^ One study reported a PTSD-hypertension association in younger women^52^ but did not examine ASCVD. However, it is difficult to compare studies of PTSD and CVD among women because of differences in study population (e.g., active-duty in this study vs. veteran or civilian women; sample age);^53–55^ endpoints studied (e.g., single^14,52^ vs. composite CVD diagnoses^16,56–59^); PTSD assessment method (diagnostic codes vs. questionnaire/interview);^14–16^ and follow-up length.^48,52^

### Metabolic Risk Factors, Depression, and Sleep Disorders

Just as PTSD was associated with CVD independent of comorbidities, as in prior studies,^19,21,22,60^ we found that MRF (obesity, T2DM), BRF (tobacco and alcohol use disorders), depression, and sleep disorders were each associated with hypertension and ASCVD, independent of PTSD and of each other.^61^ Adjusting for comorbidities attenuated the strength of PTSD associations with CVD; this is consistent with prior evidence that the presence of disorders associated with PTSD (e.g., T2DM) attenuates the association between PTSD and CVD.^62,63^ This effect likely reflects shared variance between PTSD and comorbidities and overlapping mechanisms of disease.^10, 12, 27, 28, 32–34^

A novel finding is that comorbid conditions modify the relationship between PTSD and hypertension, but not ASCVD; specifically, associations between PTSD and hypertension differed between those with vs. without MRF, depression, and sleep disorders. Results further suggest a cumulative effect of PTSD and comorbidities on standardized hypertension risk. Specifically, absolute risk of hypertension is consistently highest among those with both PTSD and a comorbidity, lowest without PTSD or comorbidity, and fall between those values for individuals with either PTSD or a comorbidity. However, significant interactions between PTSD and comorbid disorders and between PTSD and gender were only found for hypertension and not ASCVD. These results may be partly explained by our predominantly healthy and young population, the relatively low prevalence of ASCVD at younger ages, and a higher likelihood of clinically evident hypertension prior to middle age.^42,43^ Moreover, hypertension may partially mediate the relationship between PTSD and ASCVD, as indicated by the finding that with hypertension included as a covariate in the full ASCVD model, hypertension was associated with the ASCVD outcome, but PTSD was no longer a significant predictor.

Possible mechanisms involved in the direct and interactive effects of PTSD and comorbidities on CVD include PTSD- and stress-related disruptions in HPA axis and other neuroendocrine and metabolic processes; alterations in CNS and autonomic activity; inflammatory/immune changes; and biological effects of sleep disruption.^10,12,27,28,32–34,37,60,64–67^ These physiological processes are associated with PTSD and comorbidities,^37,66^ and there are interactions among these processes, including autonomic nervous system, HPA axis, and inflammatory processes,^66,68^ and additive and/or synergistic relationships among CVD risk factors (e.g., diabetes, smoking, alcohol use).^61,69^ Further research is needed to understand shared disease mechanisms that account for the present findings.

### Study Strengths and Limitations

Unlike civilian healthcare, the military healthcare system serves and maintains health and administrative databases for all active-duty soldiers, regardless of socioeconomic level, race, or gender. This contrasts with the Department of Veterans Affairs (VA) system, since some veterans routinely seek care outside the VA. Although our large active-duty enlisted population may increase the likelihood that findings are generalizable to younger, otherwise healthy individuals, selection bias and a relatively low percentage of female soldiers in this sample may also limit generalizability of our findings.

Limitations are also inherent in using diagnostic codes as endpoints. Diagnostic codes may vary between clinicians and are available only if individuals seek care. Some servicemembers with PTSD or other mental disorders believe they do not need treatment or avoid seeking professional help because of stigma;^70^ therefore, it is possible that PTSD and/or depression might have been present but undiagnosed before CVD diagnosis.

As noted previously, associations between gender and ASCVD may not be evident in this younger and healthy cohort. Moreover, the present findings may only generalize to the diagnoses used to quantify hypertension (initial hypertension diagnosis and no current/prior ASCVD) and ASCVD (composite endpoints of initial diagnosis of atherosclerosis, MI, stroke, or congestive heart failure). However, there are several commonalities in pathological mechanisms among the diagnoses used for ASCVD outcome, and positive associations with PTSD have been demonstrated with both single ^4–6,14^ and composite^16,56–59^ CVD endpoints.^9–11,66^ Nevertheless, heterogeneity among diagnoses might have obscured interactions between comorbidities and PTSD for ASCVD.

### Conclusions and Clinical Implications

It is notable that we observed the association of PTSD with hypertension, ASCVD, and comorbidities even among relatively healthy, predominantly young soldiers. Thus, physical health consequences of PTSD may begin early, develop subclinically, and have cumulative, long-term effects on morbidity/mortality, quality of life, and the healthcare system.

PTSD is currently defined and treated as a mental health disorder,^1^ perhaps because its psychiatric symptoms are overt, debilitating, and readily tied to trauma.^11,71^ However, evidence indicates that PTSD symptoms are part of a broader systemic disorder with wide-ranging biological and behavioral elements.^19,21,60,72^ The present study reveals important associations and interactions among PTSD, comorbidities, and risk of CVD, and supports a systemic understanding of PTSD.

Current evidence is inconclusive regarding whether treating PTSD using traditional approaches reduces risk of CVD or other comorbid disorders.^67,73^ Empirically-validated psychological/psychiatric treatments for PTSD address and reduce psychological symptomatology, but physical comorbidities associated with PTSD warrant attention from a range of healthcare specialties.^19,67,73^ Thus, the present findings have clinical relevance for identifying individuals at higher risk of CVD, and highlight the need to implement multidisciplinary treatment approaches that address both psychological symptoms of PTSD and associated comorbidities.

## Data Availability

All data produced in the present study are available upon approval of the Army Study to Assess Risk and Resilience in Servicemembers (Army STARRS) and the U.S. Department of Defense. The data are not publicly
available due to privacy or ethical restrictions.

## ACKNOWLEDGEMENTS

The Army STARRS Team consists of Co-Principal Investigators: Robert J. Ursano, MD (USUHS) and Murray B. Stein, MD, MPH (University of California San Diego and VA San Diego Healthcare System). <u>Site Principal Investigators</u>: James Wagner, PhD (University of Michigan) and Ronald C. Kessler, PhD (Harvard Medical School). <u>Army scientific</u> <u>consultant/liaison</u>: Kenneth Cox, MD, MPH (Office of the Assistant Secretary of the Army (Manpower and Reserve Affairs). <u>Other team members</u>: Pablo A. Aliaga, MS (USUHS); David M. Benedek, MD (USUHS); Laura Campbell-Sills, PhD (University of California San Diego); Carol S. Fullerton, PhD (USUHS); Nancy Gebler, MA (University of Michigan); Meredith House, BA (University of Michigan); Paul E. Hurwitz, MPH (Uniformed Services University); Sonia Jain, PhD (University of California San Diego); Tzu-Cheg Kao, PhD (USUHS); Lisa Lewandowski-Romps, PhD (University of Michigan); Alex Luedtke, PhD (University of Washington and Fred Hutchinson Cancer Research Center); Holly Herberman Mash, PhD (USUHS); James A. Naifeh, PhD (USUHS); Matthew K. Nock, PhD (Harvard University); Nur Hani Zainal, PhD (Harvard Medical School); Nancy A. Sampson, BA (Harvard Medical School); and Alan M. Zaslavsky, PhD (Harvard Medical School).

## Disclaimers

The opinions and assertions expressed herein are those of the authors and do not reflect the official policy or position of the Uniformed Services University of the Health Sciences or the Department of Defense. The contents of this publication also are the sole responsibility of the authors and do not necessarily reflect the views, opinions or policies of The Henry M. Jackson Foundation for the Advancement of Military Medicine, Inc. Mention of trade names, commercial products, or organizations does not imply endorsement by the U.S. Government. The contents are solely the responsibility of the authors and do not necessarily represent the views of the NIMH, the Department of the Army, the Department of Defense, or the Department of Veteran Affairs.

