## Supplement e-tables 1-7 for "PTSD, Comorbidities, Gender, and Increased Risk of Cardiovascular Disease in a Large Military Cohort"

**SUPPLEMENT: eTables 1-7**

**Supplement eTable 1**

*List and Brief Descriptions of Administrative Data Systems in the Army STARRS Historical Administrative Data Study (HADS) Included in the Current Study*

| Database Acronym | Description |
|---|---|
| DMDC/CTS | DEFENSE MANPOWER DATA CENTER (DMDC) / CONTINGENCY TRACKING SYSTEM (CTS): Collection of activation, mobilization, and deployment data. Provides information to DoD decision makers and includes a CTS Deployment File used for tracking the location of deployed personnel. |
| DMDC/Master Personnel & DMDC/Transaction files | DEFENSE MANPOWER DATA CENTER (DMDC) / MASTER PERSONNEL & TRANSACTION FILES: The Active-duty Master File provides an inventory of all individuals on active-duty (excluding reservists on active-duty for training) at a point in time. It is a standardized and centralized database of present and past members of the active-duty force. Personal data elements include social security number, education level, home of record, date of birth, marital status, number of dependents, race, ethnic group, and name. Military data elements include Service, pay grade, Armed Forces Qualification Test percentile (enlisted only), source of commission (officers only), military primary duty and secondary occupation, Unit Identification Code, months of service, duty location, Estimated Termination of Service date, basic active service date, date of current rank, pay entry base date, foreign language ability, and major command code. |
| MDR | MILITARY HEALTH SYSTEM DATA REPOSITORY (MDR): This database contains information about medical, dental, pharmaceutical, and ancillary claims data for both in network and purchased care as well as both inpatient and outpatient treatment. Data are collected on both Army personnel and their beneficiaries. |
| TMDS | THEATER MEDICAL DATA STORE (TMDS): Used to track, analyze, view and manage Soldier medical treatment information recorded in the theater of operations. Features of TMDS: accessibility and visibility of servicemembers' deployed medical records, outpatient and inpatient treatment records created in theater facilities, treatment records from other applications, reports on movement of patients, patient status and injury/illnesses. |
| TRAC2ES | TRANSCOM REGULATING AND COMMAND & CONTROL EVACUATION SYSTEM (TRAC2ES): A tracking system for all medical transfers across the world for all DOD services. |

**Supplement eTable 2**

*International Classification of Diseases, Ninth Revision–Clinical Modification (ICD-9-CM) Codes Used to Identify Cardiovascular Disease*

| Cardiac Grouping | Severity[a] | Number of Soldiers[b] | Included Diagnoses | ICD-9-CM Codes |
|---|---|---|---|---|
| Hypertension | 5 | 49,656 | Malignant Hypertension | 401.00 |
| | | | Benign Hypertension | 401.10 |
| | | | Hypertension, Unspecified | 401.90 |
| | | | Hypertensive Heart Disease | 402 |
| | | | Essential Hypertension | 410 |
| Coronary Artery Disease | 4 | 1,335 | Angina Pectoris | 413 |
| | | | Coronary Artery Disease | 414.00 |
| | | | Coronary Atherosclerosis | 414.40 |
| | | | Cardiomyopathy | 425 |
| | | | Cardiovascular Disease, Unspecified | 429.20 |
| | | | Peripheral Vascular Disease | 433.90 |
| | | | Atherosclerosis | 440.00 |
| Stroke | 3 | 516 | Occlusion and Stenosis | |
| | | | of Basilar Artery with Cerebral Infarction | 433.01 |
| | | | of Carotid Artery with Cerebral Infarction | 433.11 |
| | | | of Vertebral Artery with Cerebral Infarction | 433.21 |
| | | | of Multiple/Bilateral Precerebral Arteries with Cerebral Infarction | 433.31 |
| | | | of Other Precerebral Artery with Cerebral Infarction | 433.81 |
| | | | of Unspecified Precerebral Artery with Cerebral Infarction | 433.91 |
| | | | Cerebral Thrombosis with Cerebral Infarction | 434.01 |
| | | | Cerebral Embolism with Cerebral Infarction | 434.11 |
| | | | Cerebral Artery Occlusion with Cerebral Infarction | 434.91 |
| Myocardial Infarction | 2 | 87 | Myocardial Infarction, Acute, Anterolateral | 410.00 |
| | | | Myocardial Infarction, Acute, Anterior | 410.10 |
| Congestive Heart Failure | 1 | 489 | Congestive Heart Failure | 428.00 |

[a] From least (5) to most (1) severe.

[b] Number of soldiers with this diagnosis as their most severe

**Supplement eTable 3**

*International Classification of Diseases, Ninth Revision–Clinical Modification (ICD-9-CM) Codes Used to Identify Risk Factors*

| Risk Factor Grouping | Included Diagnoses | ICD-9-CM Codes |
|---|---|---|
| PTSD | PTSD | 309.81 |
| Metabolic risk factors | Diabetes Mellitus, II Controlled | 250.00 |
| | Diabetes Mellitus, II Uncontrolled | 250.02 |
| | Metabolic Syndrome | 277.70 |
| | Overweight and Obesity | 278.00 |
| Behavioral risk factors | Alcohol Withdrawal Delirium | 291.00 |
| | Alcohol-Induced Persistent Amnestic Disorder | 291.10 |
| | Alcohol-Induced Persisting Dementia | 291.20 |
| | Alcohol-Induced Psych Disorder with Hallucinations | 291.30 |
| | Idiosyncratic Alcohol Intoxication | 291.40 |
| | Alcohol-Induced Psych Disorder with Delusions | 291.50 |
| | Alcoholic Psychosis NEC | 291.80 |
| | Alcohol Withdrawal | 291.81 |
| | Alcohol Induced Sleep Disorder | 291.82 |
| | Other Specified Alcohol-Induced Mental Disorder | 291.89 |
| | Unspecified Alcohol-Induced Mental Disorder | 291.90 |
| | Alcohol Intoxication – Unspecified | 303.00 |
| | Alcohol Intoxication – Continued | 303.01 |
| | Alcohol Intoxication – Episodic | 303.02 |
| | Alcohol Intoxication – In Remission | 303.03 |
| | Alcohol Dependence NEC/NOS | 303.90 |
| | Alcohol Dependence NEC/NOS – Continuous | 303.91 |
| | Alcohol Dependence NEC/NOS – Episodic | 303.92 |
| | Alcohol Dependence NEC/NOS – In Remission | 303.93 |
| | Alcohol Abuse | 305.00 |
| | Alcohol Abuse – Unspecified | 305.00 |
| | Alcohol Abuse – Continuous | 305.01 |
| | Alcohol Abuse – Episodic | 305.02 |
| | Alcohol Abuse – In Remission | 305.03 |
| | Tobacco Use Disorder | 305.1 – 305.13 |

| | | |
|---|---|---|
| Depressive disorders | Major Depressive Disorder, Single Episode | 296.20 – 296.29 |
| | Major Depressive Disorder, Recurrent Episode | 296.30 – 296.39 |
| | Atypical Depressive Disorder | 296.82 |
| | Unspecified Episodic Mood Disorder | 296.90 |
| | Other Specified Episodic Mood Disorder | 296.99 |
| | Dysthymic Disorder | 300.40 |
| | Neurasthenia | 300.50 |
| | Adjustment Disorder with Depressed Mood | 309.00 |
| | Prolonged Depressive Reaction | 309.10 |
| | Depressive Disorder NOS | 311.00 |
| | Misery and Unhappiness Disorder | 313.10 |
| Sleep disorders | Nonorganic Sleep Disorder, NOS | 307.40 |
| | Transient Insomnia | 307.41 |
| | Persistent Insomnia | 307.42 |
| | Transient Hypersomnia | 307.43 |
| | Persistent Hypersomnia | 307.44 |
| | Circadian Rhythm Sleep Disorder, Nonorganic | 307.45 |
| | Sleep Arousal Disorder | 307.46 |
| | Sleep Stage Dysfunction, NOS | 307.47 |
| | Repetitive Intrusions of Sleep | 307.48 |
| | Nonorganic Sleep Disorder, Other | 307.49 |
| | Sleep Disorder/Disturbance | 780.50 |
| | Insomnia with Sleep Apnea | 780.51 |
| | Insomnia, Unspecified | 780.52 |
| | Hypersomnia with Sleep Apnea | 780.53 |
| | Hypersomnia, Unspecified | 780.54 |
| | Disruptions of 24-Hour Sleep-Wake Cycle | 780.55 |
| | Dysfunctions of Sleep Stages or Arousal | 780.56 |
| | Unspecified Sleep Apnea | 780.57 |
| | Sleep Related Movement Disorder, NOS | 780.58 |
| | Insomnia, Other | 780.59 |

Note. NEC = Not Elsewhere Classifiable; NOS = Not Otherwise Specified

**Supplement eTable 4**

*Overall Crude Rates of Hypertension Only and Arteriosclerotic Cardiovascular Disease per 1,000 Person-Years by Posttraumatic Stress Disorder Diagnosis, Gender, and Age Group*

| | Cases | Total | Crude Rate | Cases | Total | Crude Rate |
|---|---|---|---|---|---|---|
| PTSD | | | | | | |
| No | 47,455 | 29,429,948 | 19.35 | 2,320 | 30,377,118 | 0.92 |
| Yes | 2,201 | 442,238 | 59.72 | 107 | 508,460 | 2.53 |
| Total | 49,656 | 29,872,186 | 19.95 | 2,427 | 30,885,578 | 0.94 |
| Gender | | | | | | |
| Women | 5,638 | 4,130,300 | 16.38 | 335 | 4,247,832 | 0.95 |
| Men | 44,018 | 25,741,886 | 20.52 | 2,092 | 26,637,746 | 0.94 |
| Total | 49,656 | 29,872,186 | 19.95 | 2,427 | 30,885,578 | 0.94 |
| Age | | | | | | |
| ≤ 20 years | 2,241 | 4,629,144 | 5.81 | 112 | 4,634,667 | 0.29 |
| 21-24 years | 8,692 | 9,168,494 | 11.38 | 253 | 9,311,001 | 0.33 |
| 25-29 years | 10,011 | 7,006,278 | 17.15 | 263 | 7,246,099 | 0.44 |
| 30-34 years | 8,652 | 4,056,296 | 25.60 | 305 | 4,222,457 | 0.87 |
| 35-39 years | 10,262 | 3,108,310 | 39.62 | 512 | 3,362,736 | 1.83 |
| ≥ 40 years | 9,798 | 1,903,665 | 61.76 | 982 | 2,108,618 | 5.59 |
| Total | 46,656 | 29,872,186 | 19.95 | 2,427 | 30,885,578 | 0.94 |

Note. ASCVD = arteriosclerotic cardiovascular disease (coronary artery disease, stroke, myocardial infarction, congestive heart failure); PTSD = posttraumatic stress disorder

**Supplement eTable 5**

*Crude Rates of Hypertension Only and Arteriosclerotic Cardiovascular Disease per 1,000 Person-Years by Posttraumatic Stress Disorder Diagnosis in Women and Men*

| | | Hypertension Only | | | Arteriosclerotic CVD | | |
|---|---|---|---|---|---|---|---|
| | PTSD | Cases | Total | Crude Rate | Cases | Total | Crude Rate |
| Women | No | 5,475 | 4,066,208 | 16.16 | 321 | 4,179,748 | 0.92 |
| | Yes | 163 | 64,092 | 30.52 | 14 | 68,084 | 2.47 |
| Men | No | 41,980 | 25,363,740 | 19.86 | 1,999 | 26,197,370 | 0.92 |
| | Yes | 2,038 | 378,146 | 64.67 | 93 | 440,376 | 2.53 |
| Total | | 49,656 | 29,872,186 | 19.95 | 2,427 | 30,885,578 | 0.94 |

Note. CVD = cardiovascular disease; PTSD = posttraumatic stress disorder

**Supplement eTable 6**

*Crude Rates of Hypertension Only and Arteriosclerotic Cardiovascular Disease per 1,000 Person-Years by Posttraumatic Stress Disorder Diagnosis in Six Age Groups*

| | | Hypertension Only | | | Arteriosclerotic CVD | | |
|---|---|---|---|---|---|---|---|
| | PTSD | Cases | Total | Crude Rate | Cases | Total | Crude Rate |
| ≤ 20 years | No | 2,218 | 4,618,203 | 5.76 | 110 | 4,625,251 | 0.29 |
| | Yes | 23 | 10,940 | 25.23 | 2 | 9,416 | 2.55 |
| 21-24 years | No | 8,325 | 9,059,783 | 11.03 | 244 | 9,203,093 | 0.32 |
| | Yes | 367 | 108,710 | 40.51 | 9 | 107,907 | 1.00 |
| 25-29 years | No | 9,387 | 6,862,326 | 16.41 | 247 | 7,081,339 | 0.42 |
| | Yes | 624 | 143,951 | 52.02 | 16 | 164,760 | 1.17 |
| 30-34 years | No | 8,205 | 3,975,320 | 24.77 | 292 | 4,117,442 | 0.85 |
| | Yes | 447 | 80,976 | 66.24 | 13 | 105,015 | 1.49 |
| 35-39 years | No | 9,841 | 3,048,869 | 38.73 | 493 | 3,284,871 | 1.80 |
| | Yes | 421 | 59,442 | 84.99 | 19 | 77,865 | 2.93 |
| ≥ 40 years | No | 9,479 | 1,865,446 | 60.98 | 934 | 2,065,121 | 5.43 |
| | Yes | 319 | 38,218 | 100.16 | 48 | 43,497 | 13.24 |
| Total | | 49,656 | 29,872,186 | 19.95 | 2,427 | 30,885,578 | 0.94 |

**Supplement eTable 7**

*Associations of First PTSD Diagnosis with Subsequent ASCVD Diagnosis Controlling for Hypertension in a Full Multivariable Model Among Regular Army Enlisted SoldiersI*[a,b,c,d]

| | OR | 95% CI | $\chi^2$, *p* |
|---|---|---|---|
| Gender | | | $\chi^2_1$=1.4, *p*=0.2315 |
| Female | 0.9 | [0.82, 1.05] | |
| Male | 1.0 | – | |
| Age (years) | | | $\chi^2_5$=972.7, *p*<0.0001* |
| ≤ 20 | 0.3 | [0.24, 0.40] | |
| 21–24 | 0.4 | [0.32, 0.47] | |
| 25–29 | 0.5 | [0.42, 0.60] | |
| 30–34 | 1.0 | – | |
| 35–39 | 2.0 | [1.69, 2.27] | |
| ≥ 40 | 5.1 | [4.37, 5.86] | |
| Race | | | $\chi^2_4$=1.5, *p*=0.8278 |
| White Non-Hispanic | 1.0 | – | |
| Black | 1.0 | [0.94, 1.14] | |
| Hispanic | 1.0 | [0.84, 1.12] | |
| Asian | 0.9 | [0.75, 1.17] | |
| Other | 1.1 | [0.84, 1.36] | |
| Time in service (years) | | | $\chi^2_3$=6.9, *p*=0.0754 |
| 1–2 | 0.8 | [0.64, 1.08] | |
| 3–4 | 1.0 | [0.83, 1.20] | |
| 5–10 | 1.0 | – | |
| ≥ 10 | 0.9 | [0.74, 0.99] | |
| Rank | | | $\chi^2_4$=15.1, *p*=0.0045 |
| E1–E2 | 1.6 | [1.26, 2.13] | |
| E3 | 1.2 | [0.94, 1.56] | |
| E4 | 1.0 | – | |
| E5–E6 | 1.0 | [0.81, 1.14] | |
| E7–E9 | 0.9 | [0.77, 1.15] | |
| Deployed[f] | | | $\chi^2_1$=35.3, *p*<0.0001* |
| Yes | 0.8 | [0.70, 0.83] | |
| No | 1.0 | – | |

| | | | |
|---|---|---|---|
| MRF | | | $\chi^2_1$=14.4, *p*=0.0001* |
| Yes | 1.3 | [1.13, 1.46] | |
| No | 1.0 | – | |
| BRF | | | $\chi^2_1$=122.3, *p*<0.0001* |
| Yes | 1.8 | [1.59, 1.94] | |
| No | 1.0 | – | |
| Depression | | | $\chi^2_1$=9.7, *p*=0.0018 |
| Yes | 1.2 | [1.08, 1.41] | |
| No | 1.0 | – | |
| Sleep disorders | | | $\chi^2_1$=21.6, *p*<0.0001* |
| Yes | 1.4 | [1.20, 1.55] | |
| No | 1.0 | – | |
| Hypertension | | | $\chi^2_1$=592.2, *p*<0.0001* |
| Yes | 3.3 | [3.00, 3.63] | |
| No | 1.0 | – | |
| PTSD | | | $\chi^2_1$=7.8, *p*=0.0052 |
| Yes | 1.4 | [1.10, 1.68] | |
| No | 1.0 | – | |

Note. ASCVD = arteriosclerotic CVD; CVD = cardiovascular disease; MRF = medical risk factors; BRF = behavioral risk factors; HADS = Army STARRS Historical Administrative Data Study; PTSD = posttraumatic stress disorder; STARRS = Army Study to Assess Risk and Resilience in Servicemembers.

*$p < 0.00167$ (sample size-adjusted alpha level).

[a] This sample of Regular Army enlisted soldiers were on active-duty at any point during 2004-2009 and is a subset of the total Army STARRS HADS.

[b] The sample included 2,427 ASCVD cases and 87,722 unweighted control person-months. All control person-months were assigned a weight of 362.1 to adjust for under-sampling.

[d] Multivariate model includes all of the variables listed plus historical time (using a dummy predictor variable for calendar month and year).
